# Mental health challenges among adolescents and young adults with perinatally acquired HIV: key findings from the I’mPossible Program in India

**DOI:** 10.1101/2025.03.20.25324329

**Authors:** Ashley A. Sharma, Michael Babu Raj, Babu Seenappa, Siddha Sannigrahi, Kacie Filian, Esha Nobbay, Suhas Reddy, Prashant Laxmikanth, Sanya Thomas, Aastha Kant, SK Satish Kumar, Sunil S. Solomon, Lakshmi Ganapathi, Anita Shet

## Abstract

Adolescents and young adults with HIV are reported to be at high risk for common mental health disorders (CMD), but studies in India are limited. The risks may be greater among adolescents and young adults with perinatally acquired HIV (APHIV), as they face lifelong medical challenges, higher levels of stigma, and stressors related to disclosure, adherence, and transition to adult care. We screened for depression and generalized anxiety disorder (GAD) and explored psychosocial experiences among a cohort of APHIV in southern India to inform development of tailored CMD interventions. Between March-June 2023, we administered a cross-sectional survey to participants in the I’mPossible Fellowship, a peer-led mentorship program for APHIV in southern India. Survey design and administration incorporated participatory research principles, wherein trained peer mentors (youth investigators) administered screening tools for depression (Patient Health Questionnaire-9: PHQ-9), anxiety (Generalized Anxiety Disorder-7: GAD-7), resilience (Child Youth Resilience Measure-Revised – CYRM-R), and an abbreviated HIV stigma Scale. Subsequently we conducted focus group discussions with selected participants to explore perspectives on mental health, stigma and perceived pathways towards improved health outcomes. We used multivariable regression to identify correlates of positive CMD screens and inductively analyzed focus group transcripts. Among 185 APHIV survey participants, mean age was 18.6 years (SD 3.5 years); 63.2% were male. Most (91.9%) had lost one or both parents, and 43.2% lived in child care institutions (CCIs). The majority (90.4%) were virally suppressed (VL<150 copies/mL). PHQ-9 and GAD-7 scores in the severity category of mild or above were defined as positive. A high proportion screened positive for at least one CMD (62.7%), depression alone (25.9%), GAD alone (7%), or both (29.7%). Externalized stigma was high (74.6%), reinforcing disclosure concerns (81.1%). Loss of both parents was associated with increased odds of anxiety (aOR 2.10, 95% CI 1.07-4.09). Exploration of anxiety and depression-related factors among APHIV revealed themes across the socioecological model (SEM) constructs that included uncertainty about transitioning to adult care, ART adherence challenges, and maladaptive coping mechanisms. Family support, disclosure fears, school pressures, stigma, and evolving societal attitudes also shaped participants’ mental health experiences. The significant burden of positive screens for CMD among APHIV requires HIV programs in India to prioritize youth-tailored stigma-informed mental health interventions alongside strategies for successful adult care transition and long-term viral suppression.

## Introduction

As of 2022, 1.65 million adolescents aged 10-19 years and 3.1 million young people aged 15-24 years were living with HIV worldwide.^1,2^ Collectively, adolescents and young adults (defined as those between the ages 10-24 years, referred to as ‘youth’ in this paper) constitute at least a third of all new HIV infections globally.^2,3 4^ Adolescence is a distinct developmental phase characterized by significant physical, psychological, and social changes, marking a critical transition from childhood to adulthood.^5,6^ This period is associated with both opportunities for social and emotional growth, as well as a myriad of social, emotional, and behavioral challenges.^7^ Approximately 50% of mental health disorders are estimated to have onset during adolescence,^8,9^ and they contribute to suicide, a leading cause of death among adolescents in all regions of the world.^10–12^ Multiple studies indicate that young people with HIV, particularly adolescents and young adults with perinatally acquired HIV (APHIV) have a high prevalence of common mental health disorders (CMD) such as depression, generalized anxiety disorder (GAD), and post-traumatic stress disorder (PTSD).^13–16^ Systematic reviews estimate that 26.1% of youth with HIV have depression,^15^ while 45.6% have anxiety.^17^ The burden of CMD is higher among this population compared to youth without HIV and is influenced by distinct physiological and psychosocial vulnerabilities, including the physiological effects of HIV and treatment regimens, the burden of managing a chronic illness,^18^ and psychosocial stressors such as pervasive stigma and discrimination.^19–22^

CMD have a significant impact on HIV treatment outcomes; co-existing unaddressed depression and/or anxiety disorders are associated with decreased antiretroviral therapy (ART) adherence, increased sexual risk behaviors, lower viral suppression, higher attrition across the HIV treatment cascade, and increased mortality rates.^23–26^ While ART has contributed to decreased HIV-related mortality overall,^27–29^ this double burden of the HIV epidemic and CMD predominantly affects low- and middle-income countries (LMICs), where approximately 90% of APHIV live.^30,31^ Studies evaluating CMD among APHIV in LMICs have primarily focused on countries in sub-Saharan Africa and to a lesser extent, in Southeast Asia. Such research is limited in India^32,33^ although a substantially large APHIV population live in India, with numbers comparable to countries in sub-Saharan Africa.^34–36^

Globally there is a paucity of youth-tailored evidence-based mental health interventions for CMD among APHIV.^37^ More recently interventions spanning family-based psychosocial and economic interventions, as well as group and community-based interventions have been developed in pilot studies and randomized controlled trials among youth with HIV.^37,38^ Some of these interventions include elements of cognitive-behavioral therapy (CBT), mindfulness, problem-solving therapy, and other evidence-based mental health interventions.^39–42^ In India’s context, the lack of well-studied mental health interventions specifically tailored for APHIV is stark. There is a great need for interventions for preventing and addressing CMD to be integrated into public sector HIV treatment programs, where most APHIV seek care.

In 2021, we established the I’mPossible Fellowship, an 18-month program in which young adults living with HIV, termed ‘fellows’ receive training and supervision to provide support related to ART adherence, education, and psychosocial needs to children and adolescents with HIV, termed ‘peers’, through one-on-one mentorship. While peer support can enhance HIV and broader psychosocial outcomes,^39–42^ addressing specific CMD necessitates data-informed contextual evidence-based interventions. In this study, we aimed to screen for CMD among APHIV participants of the I’mPossible Fellowship in India, explore their psychosocial experiences, and understand structural and social determinants of CMD to inform the development of youth-tailored mental health interventions.

## Methods

### Participants and study setting

The I’mPossible Fellowship program was established in the southern states of Karnataka and Tamil Nadu through collaborative efforts with community-based organizations providing integral support, such as safe residential care, medical treatment, education, and psychosocial assistance, to children and APHIV. Eligible participants in this study were between ages 15-24 years, received a diagnosis of HIV prior to 10 years of age, were resident in child care institutions (CCIs) or in family-based care. Interventions integral to the I’mPossible Fellowship program are described in detail elsewhere.^43^ In addition to the regular structured interventions within the program, CCI supervisors were available to provide oversight and facilitate referrals to state welfare schemes, pediatric specialists, psychiatrists, and other mental health professionals. Although it provides well-rounded support to APHIV, the I’mPossible program in its current form does not incorporate evidence-based interventions to address CMD among peers, and higher level needs such as treatment of CMD are primarily addressed through referrals. Exploring CMD and psychosocial experiences in this study was a crucial step towards developing targeted interventions addressing mental health in this population.

We conducted the study between 17 April 2023 and 24 November 2023. Participants included peers from the first cohort of the I’mPossible Fellowship that comprised 257 peers. Those with ages 13 years and above were included in this study. All had a diagnosis of HIV prior to age 10 years (definition of perinatally-acquired HIV) and were aware of their HIV status. For participants with age ≥18 years oral, informed consent was obtained by trained research staff, and for participants <18 years, oral permission from a parent or legal guardian followed by oral assent from the minor participant was obtained.

### Youth investigators and community-based participatory research

For survey development, adaptation, and administration, we incorporated a community-based participatory approach by including 5 fellows from the second cohort and 4 fellows from third cohort of the I’mPossible Fellowship (who had not directly provided mentorship to study participants).^44^ These ‘youth investigators’ actively shaped the research process by culturally adapting the English, Tamil, and Kannada versions of CMD screening instruments utilized in the survey through iterative discussions with experts, while simultaneously drawing on their own perspectives as APHIV. Prior to conducting study assessments, they also underwent certification in human subjects’ protection, training in survey administration techniques, and mental health screening, including recognizing when peer participants needed referrals to counselors or mental health professionals for further evaluation and care.

### Mental Health Surveys

The comprehensive mental health survey comprised a general questionnaire and specific instruments to measure CMD, resilience, and stigma; each of these components were previously validated in several low-and-middle income countries, including India.^45–48^ The survey components are described below:

(i) The general questionnaire collected the following data: sociodemographic characteristics, such as age, gender, place of residence; education characteristics, including current school enrollment status, reasons for school discontinuation; employment characteristics, including current employment status and workplace challenges; and HIV treatment characteristics, such as self-reported ART adherence, and viral load measurements taken within the past year.
(ii) The Child and Youth Resilience Measurement (CYRM-R).^49^ This screening tool was designed to identify strengths and resources possessed by participants, assessing the quality of peer participants’ relationships with family and caregivers, and inner strengths, such as emotional regulation and problem-solving skills.^49,50^
(iii) The Patient Health Questionnaire (PHQ-9) Questionnaire.^48,51^ To screen for depression among peer participants, we administered the PHQ-9, a 9-item questionnaire using a Likert scale scoring system to categorize depressive symptoms over the past two weeks into five severity levels: none/minimal (score 0-4), mild (score 5-9), moderate (score 10-14), moderately severe (score 15-19), and severe (score 20-27).
(iv) The General Anxiety Disorder-7 (GAD-7) Questionnaire:^48,52^ We also administered the 7-item GAD-7 questionnaire categorizing symptoms of GAD over the past two weeks into four severity levels: minimal (score 0-4), mild (score 5-9), moderate (score 10-14), and severe (score 15-21).
(v) HIV Stigma Questionnaire: To assess peer participants’ perceptions of HIV-related stigma, we created an abbreviated 4-item version of a 12-item HIV Stigma Survey,^53^ which was previously validated among adolescents with HIV in India,^54^ and itself adapted from Berger’s validated 40-item stigma questionnaire.^55,56^ The abbreviated version was intended to obtain initial insights of HIV-related stigma and comprised four statements, representing constructs of internal stigma (personalized stigma, and negative self-image), and external stigma (concerns about disclosure, and concerns regarding public attitudes towards individuals with HIV) (Supplementary Table 1).^54,56, 57^

### Focus group discussions

We conducted three FGDs, each consisting of six participants (n=18), recruited across three sites, Belgaum, Bangalore, and Krishnagiri. Aiming to explore peers’ psychosocial experiences, we used purposive recruitment strategies to include equal representation from adolescents <18 years of age and young adults between ages 18-24 years of both genders, and from APHIV living in child care institutions and those living in family-based care. The discussion guide for the focus group discussions was developed by the research team with significant input from current and former fellows and informed by the socioecological model which recognizes the multifaceted and dynamic inter-relatedness between individual and environmental factors including family, school, community and policies/governmental agencies that impact health and behavior.^58^ Within this framework, questions were formulated to assess factors contributing to overall mental health, such as peer participants’ personal and interpersonal psychosocial experiences, including experiences within and perceptions regarding their community and the larger society, existing supports, and coping behaviors. FGDs were facilitated by a trained fellow from the third cohort, with the participation of a study investigator (with MD or PhD training) who had experience facilitating focus groups. They were conducted in a private space and had an average duration of 1 hour. Focus groups for female adolescents and young adults were conducted separately from focus groups for male adolescents and young adults. Facilitators were gender concordant with focus groups. Data saturation was reached for all major topics. The discussions’ audios were recorded and translated from Kannada/Tamil to English to allow for qualitative coding of the translated transcripts. Additionally, we conducted member checking of transcripts to ensure the accuracy of the findings by reviewing them with the peer participants.

### Data Definitions

The following data definitions were established for this study: for the resilience measure, we divided scores into quartiles such that a total CYRM-R score of ≤25^th^ percentile was defined as ‘low resilience’. For sensitivity analysis, we assessed ≤15^th^, ≤33^rd^ and ≤50^th^ percentile of the total CYRM-R score, to determine if a change in threshold materially impacted the results. The presence of a positive screen for depression was defined as a severity of mild and above (score of ≥5) according to the PHQ-9,^51^ and presence of a positive screen for GAD was defined as a severity of mild and above (score of ≥5) based on the GAD-7.^59^ A detectable viral load was defined as ≥150 copies/mL, consistent with established literature.^60,61^ Self-reported ART adherence in the past month was categorized as optimal (0-<7 days of medications missed) or suboptimal (≥7 days of medications missed).

#### Data Analysis

##### Quantitative

We characterized the study cohort using summary statistics and generated multivariable logistic regression models to determine correlates of a positive screen for (1) depression alone, (2) GAD alone, and (3) at least one CMD (i.e., depression and/or anxiety). Variables included (a) sociodemographic characteristics (age, gender, place of residence); (b) education and employment characteristics (school discontinuation and employment status); (c) HIV treatment characteristics (duration on ART and viral suppression); (d) psychosocial factors (low resilience and parental deceased status). Variables assessed in univariable analysis were included in the multivariable model based on prior evidence of likely association with CMD.^8,62–72^ Analyses were performed using SPSS version 29.0 (IBM Corp, Armonk, New York).

##### Qualitative

Analysis of the FGDs’ transcripts entailed inductive and deductive approaches for thematic analysis. Initially, a coding scheme was developed through collective team discussion and insights from comparable studies. Subsequently, focus group discussions transcripts were independently coded using Dedoose software by two researchers (AAS and AK), who then convened to identify emerging themes, address discrepancies, and finalize the coding scheme. A third researcher (EN) evaluated the transcripts, overseeing the reconciliation process and ensuring consistent application of codes.

### Ethics Statement

This study obtained approval from the Institutional Review Boards of the Y.R. Gaitonde Centre for AIDS Research and Education (YRGCARE), Chennai, India and the Johns Hopkins Bloomberg School of Public Health, Baltimore, USA. Due to the combination of in-person and virtual data collection, informed consent, assent, and parental permission for this study were obtained orally and recorded through researcher-signed documentation. Written consent, assent, and permission could not be acquired due to participants’ lack of access to printers and scanners for signing forms. Institutional Review Boards approved the use of oral consent, assent, and parental permission and reviewed the forms to ensure that participants received a clear explanation of the study’s purpose, procedures, risks, and benefits

## Results

### Survey findings

#### General characteristics of participants

Among 216 age-eligible peers who were contacted for the mental health survey administration, 185 peer participants responded (12 could not be contacted, 19 cited lack of time or interest for declining). Survey participants had a mean age of 18.6 years (SD 3.5 years) and 63.2% were male, 50.3% reported that both parents were deceased, 54.6% lived in rural areas, and 43.2% lived in child care institutions (CCIs). Among those who lived in family-based care, 44.7% lived in one-parent households. Educational discontinuation was reported by 29.7%, the primary reasons being financial constraints (44.6%) and challenges in keeping up with schoolwork (23.1%). With regard to employment, 40.0% of the participants were currently working but reported facing challenges, including insufficient income (28.8%) and fears of HIV disclosure in the workplace (5.5%) (Table 1).

**Table 1.**
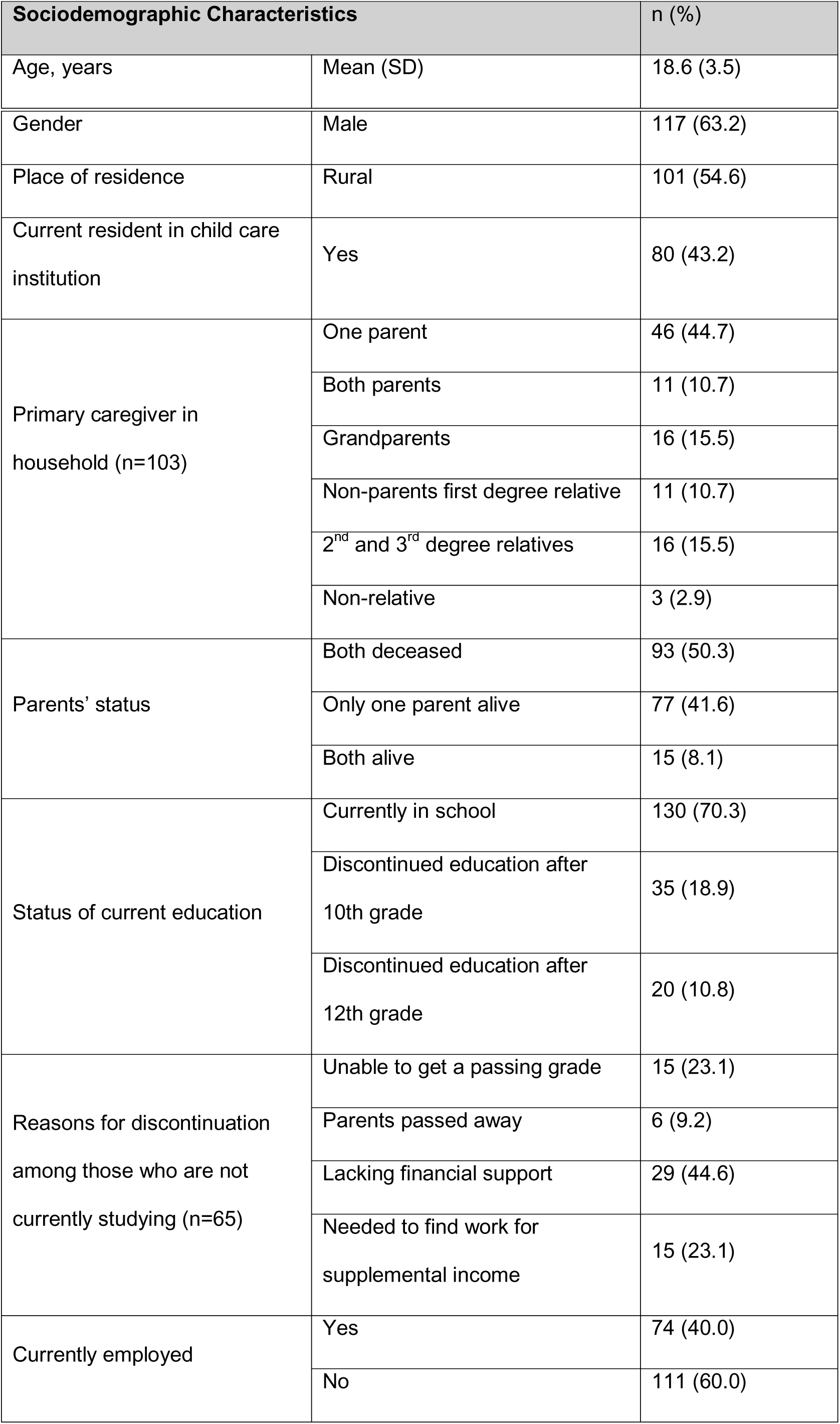
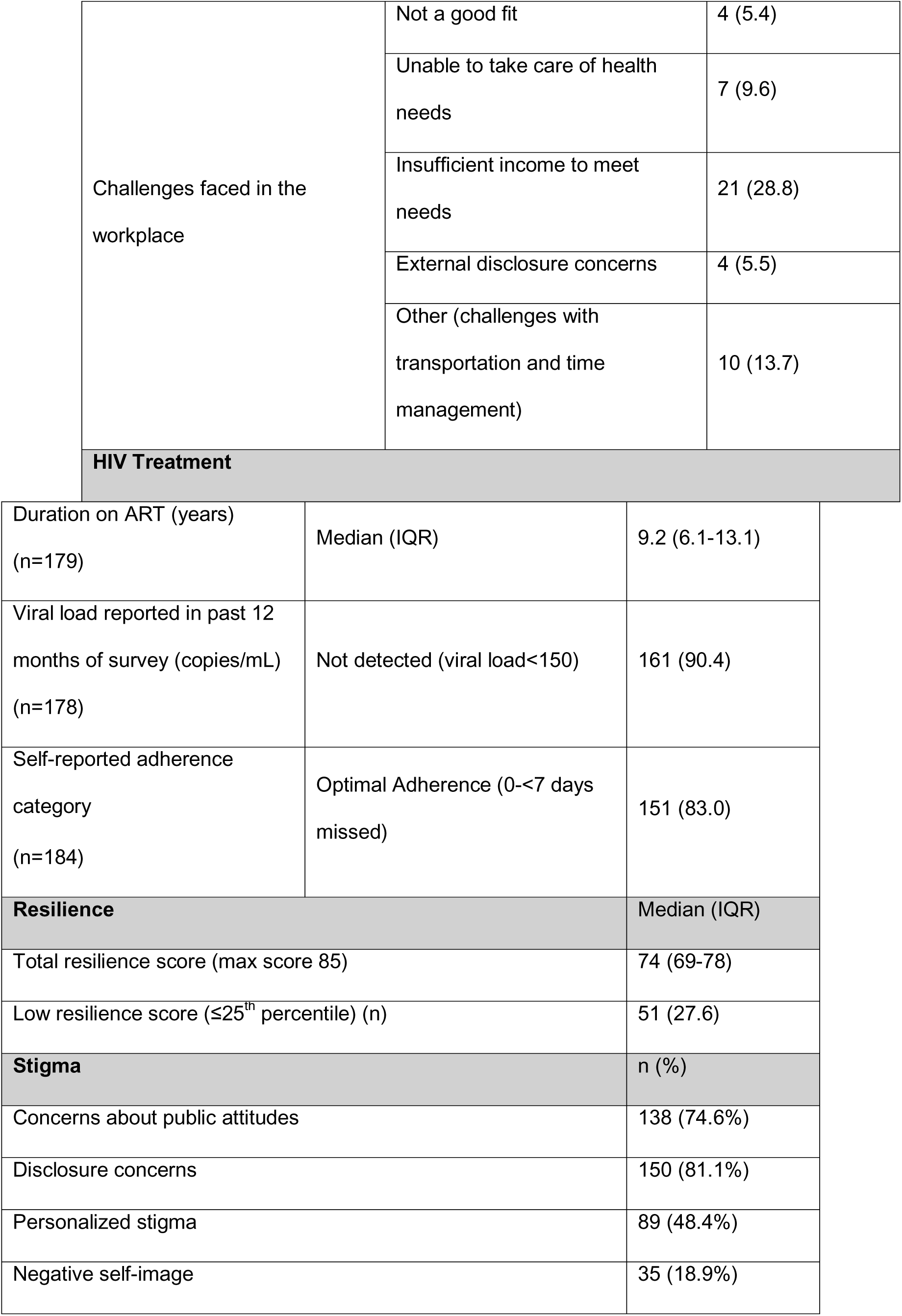
Sociodemographic Characteristics, HIV Treatment, Resilience, and Stigma Measures of AYA-PHIV participants (n=185)

| Sociodemographic Characteristics |  | n (%) |
| --- | --- | --- |
| Age, years | Mean (SD) | 18.6 (3.5) |
| Gender | Male | 117 (63.2) |
| Place of residence | Rural | 101 (54.6) |
| Current resident in child care institution | Yes | 80 (43.2) |
| Primary caregiver in household (n=103) | One parent | 46 (44.7) |
|  | Both parents | 11 (10.7) |
|  | Grandparents | 16 (15.5) |
|  | Non-parents first degree relative | 11 (10.7) |
|  | 2<sup>nd</sup> and 3<sup>rd</sup> degree relatives | 16 (15.5) |
|  | Non-relative | 3 (2.9) |
| Parents' status | Both deceased | 93 (50.3) |
|  | Only one parent alive | 77 (41.6) |
|  | Both alive | 15 (8.1) |
| Status of current education | Currently in school | 130 (70.3) |
|  | Discontinued education after 10th grade | 35 (18.9) |
|  | Discontinued education after 12th grade | 20 (10.8) |
| Reasons for discontinuation among those who are not currently studying (n=65) | Unable to get a passing grade | 15 (23.1) |
|  | Parents passed away | 6 (9.2) |
|  | Lacking financial support | 29 (44.6) |
|  | Needed to find work for supplemental income | 15 (23.1) |
| Currently employed | Yes | 74 (40.0) |
|  | No | 111 (60.0) |
| Challenges faced in the workplace | Not a good fit | 4 (5.4) |
|  | Unable to take care of health needs | 7 (9.6) |
|  | Insufficient income to meet needs | 21 (28.8) |
|  | External disclosure concerns | 4 (5.5) |
|  | Other (challenges with transportation and time management) | 10 (13.7) |
| **HIV Treatment** |  |  |
| Duration on ART (years)<br>(n=179) | Median (IQR) | 9.2 (6.1-13.1) |
| Viral load reported in past 12 months of survey (copies/mL)<br>(n=178) | Not detected (viral load<150) | 161 (90.4) |
| Self-reported adherence category<br>(n=184) | Optimal Adherence (0-<7 days missed) | 151 (83.0) |
| **Resilience** |  | Median (IQR) |
| Total resilience score (max score 85) |  | 74 (69-78) |
| Low resilience score ($\leq 25^{\text{th}}$ percentile) (n) | | 51 (27.6) |
| **Stigma** |  | n (%) |
| Concerns about public attitudes |  | 138 (74.6%) |
| Disclosure concerns |  | 150 (81.1%) |
| Personalized stigma |  | 89 (48.4%) |
| Negative self-image |  | 35 (18.9%) |

#### HIV treatment characteristics

Participants’ mean duration on ART was 9.7 years (SD 4.0 years) (Table 1). Viral suppression (viral load <150 copies/mL in the past 12 months since study recruitment) was high at 90.4%. Optimal adherence was seen among 83.0% of participants who reported <7 days of missed doses in the past month.

#### Resilience measures

Total resilience score and subscale scores were high among the peer participants (Table 1). Low resilience (CYRM-R score of ≤25th percentile) was observed in 51 participants.

#### Stigma survey

The majority of peer participants, 74.6%, reported that people with HIV face rejection when their status is disclosed, reflecting concerns about public attitudes toward individuals with HIV. Similarly, 81.1% reported making considerable efforts to keep their HIV status, or that of their parents, a secret, highlighting deep concerns about external disclosure. Nearly half, 48.4%, reported being shunned by people who became aware of their HIV status or that of their parents. However, perceptions of negative self-image were comparatively low, with 18.9% of peer participants reporting such feelings.

#### Screening for depression and generalized anxiety among APHIV

Among participants, 55.7% had a positive screen for any category of depression, and 14.6% for depression of moderate and above severity. With regard to screening results for generalized anxiety, 36.8% had a positive screen for any category of GAD and 9.8% for GAD of moderate and above severity (Table 2). Nearly two-thirds (62.7%) of participants had positive screens for either mild and above depression and/or mild and above GAD.

**Table 2.** Prevalence of positive screens for depression and general anxiety disorder among adolescents and young adults with perinatally acquired HIV (n=185)

| Common mental health disorder (CMD) | Category [scores] | n (%) |
| --- | --- | --- |
| Depression | None/minimal [0-4] | 82 (44.3) |
|  | Mild [5-9] | 76 (41.1) |
|  | Moderate [10-14] | 25 (13.5) |
|  | Moderately severe [15-19] | 2 (1.1) |
|  | Severe [20-27] | 0 (0.0) |
| Any depression | Categories mild and above [$\geq 5$] | 103 (55.7) |
| Significant depression | Categories moderate and above [$\geq 10$] | 27 (14.6) |
| Generalized Anxiety Disorder (GAD) | Minimal [0-4] | 117 (63.2) |
|  | Mild [5-9] | 50 (27.0) |
|  | Moderate [10-14] | 14 (7.6) |
| | Severe [$\geq 15$] | 4 (2.2) |
| Any anxiety | Categories mild and above [$\geq 5$] | 68 (36.8) |
| Significant anxiety | Categories moderate and above [$\geq 10$] | 18 (9.8%) |
| Screening for Common Mental Health Disorders | Both Depression and GAD | 55 (29.7) |
|  | Depression, no GAD | 48 (25.9) |
|  | GAD, no Depression | 13 (7.0) |
|  | Depression and/or GAD | 116 (62.7) |

In multivariable analysis of a positive screen for GAD (severity of mild and above; GAD-7 score of ≥ 5), double orphan status emerged as a significant correlate (aOR: 2.10, 95% CI: 1.07-4.09, p=0.030) associated with a two-fold increase in the odds of having a positive screen for GAD (Table 3). Although other correlates were not statistically significant in both univariable and multivariable models, the directionality of odds ratios suggested that female gender, school discontinuation, and current employment status could be associated with increased odds of having a positive screen for GAD. In the univariable and multivariable analysis of a positive screen for depression, no correlates showed statistically significant associations. (Table 3). However, the directionality of odds ratios in these models suggested that female gender, double orphan status, lack of viral suppression, and low resilience could be associated with increased odds of having a positive screen for depression. When at least one CMD (depression and/or GAD) was considered, none of the correlates were statistically significant in their association with having a positive screen for at least one CMD. However, the directionality of odds ratios in these models suggested that female gender, double orphan status, school discontinuation, and low resilience could be associated with increased odds of having a positive screen for at least one CMD. Low resilience (using thresholds of ≤ 15^th^, ≤ 33^rd^, and ≤ 50^th^ percentiles) in separate models did not materially change these findings.

**Table 3.** Multivariate analysis of positive screens for common mental health disorders among adolescents and young adults with perinatally acquired HIV (n=185)

|  | Generalized Anxiety Disorder |  | Depression |  | At least one common mental health disorder |  |
| --- | --- | --- | --- | --- | --- | --- |
|  | Univariate model | Multivariate model | Univariate model | Multivariate model | Univariate model | Multivariate model |
|  | OR (95% CI) | aOR (95% CI) | OR (95% CI) | aOR (95% CI) | OR (95% CI) | aOR (95% CI) |
| **Age** (continuous) | 1.02 (0.93-1.11) | 0.96 (0.85-1.08) | 1.02 (0.94-1.11) | 1.00 (0.89-1.12) | 1.03 (0.94-1.12) | 1.00 (0.89-1.12) |
| **Gender** |  |  |  |  |  |  |
| Woman | 1.64 (0.89-3.04) | 1.44 (0.72-2.91) | 1.35 (0.74-2.47) | 1.17 (0.59-2.29) | 1.56 (0.83-2.93) | 1.37 (0.68-2.75) |
| Man (Ref) |  |  |  |  |  |  |
| **Parents' status** |  |  |  |  |  |  |
| Both parents deceased | 1.73 (0.94-3.16) | 2.10 (1.07-4.09)<sup>†</sup> | 1.22 (0.68-2.17) | 1.17 (0.62-2.19) | 1.70 (0.93-3.10) | 1.69 (0.89-3.24) |
| One or both parents alive (Ref) |  |  |  |  |  |  |
| **Place or residence** |  |  |  |  |  |  |
| Rural | 1.09 (0.60-1.98) | 0.99 (0.50-1.95) | 0.82 (0.46-1.47) | 0.77 (0.40-1.49) | 0.97 (0.53-1.77) | 0.87 (0.44-1.71) |
| Urban (Ref) |  |  |  |  |  |  |
| **Living in a child care institution** |  |  |  |  |  |  |
| Yes | 0.88 (0.48-1.60) | 0.89 (0.41-1.97) | 0.80 (0.44-1.43) | 1.03 (0.49-2.19) | 0.99 (0.54-1.80) | 1.26 (0.58-2.74) |
| No (Ref) |  |  |  |  |  |  |
| **Discontinuation of school** |  |  |  |  |  |  |
| Yes | 1.51 (0.79-2.88) | 1.47 (0.68-3.18) | 1.29 (0.68-2.44) | 0.97 (0.46-2.04) | 1.33 (0.68-2.58) | 1.12 (0.52-2.40) |
| No (Ref) |  |  |  |  |  |  |
| **Currently working<sup>†</sup>** |  |  |  |  |  |  |
| Yes | 1.44 (0.70-2.96) | 1.50 (0.59-3.86) | 1.03 (0.51-2.08) | 0.74 (0.31-1.78) | 1.17 (0.57-2.40) | 0.85 (0.34-2.10) |
| No (Ref) |  |  |  |  |  |  |
| **Complete Viral Suppression (VL <150)** |  |  |  |  |  |  |
| No | 0.92 (0.32-2.61) | 1.21 (0.39-3.75) | 1.52 (0.54-4.31) | 1.14 (0.38-3.45) | 1.12 (0.39-3.18) | 0.85 (0.28-2.61) |
| Yes (Ref) |  |  |  |  |  |  |
| Duration on ART (months) | 1.00 (0.99-1.01) | 1.00 (0.99-1.01) | 1.00 (1.00-1.01) | 1.00 (0.99-1.01) | 1.00 (1.00-1.01) | 1.00 (1.00-1.01) |
| Total resilience |  |  |  |  |  |  |
| ≤ 25<sup>th</sup> centile | 1.16 (0.60-2.46) | 0.79 (0.37-1.69) | 1.88 (0.96-3.70) | 1.68 (0.81-3.52) | 1.84 (0.91-3.72) | 1.37 (0.64-2.95) |
| > 25<sup>th</sup> centile (Ref) |  |  |  |  |  |  |

### Insights from focus group discussions

Among the 18 FGD participants mean age was 16.7 years (SD = 1.5), 12 were male adolescents or young adults, 15 resided in childcare institutions, and all were studying or employed. Among the 12 FGD participants who had also completed the mental health survey, 6, 4 and 7 screened positive for depression, GAD and at least one CMD, respectively, highlighting mental health vulnerabilities within this group. Several themes emerged across the socioecological model (SEM) constructs that FGD participants identified as potential contributors or alleviators of symptoms of anxiety and/or depression. Narratives of stigma crossed multiple domains of the SEM. (Figure 1 and Table 5).

**Figure 1:**
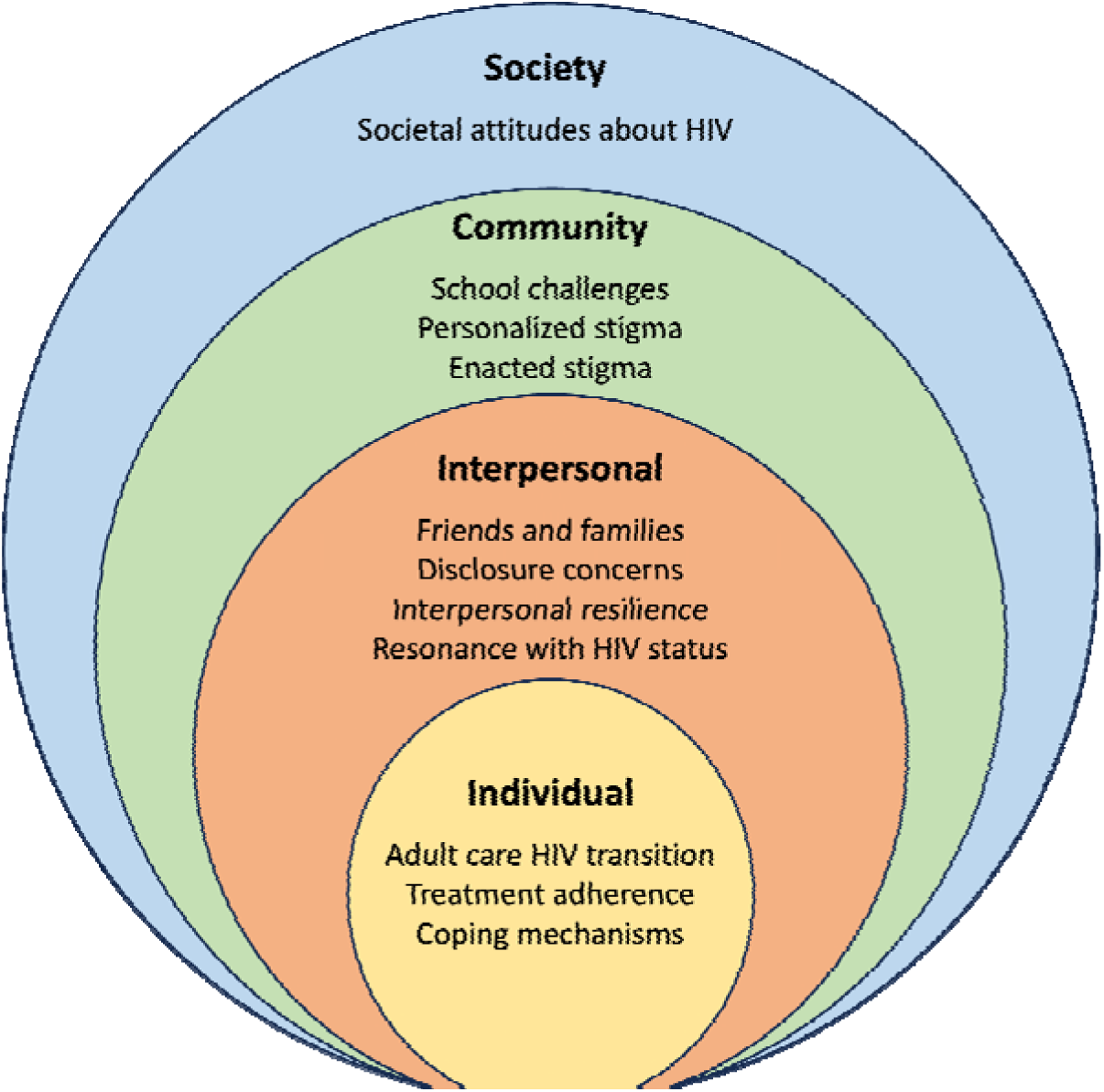
Categorization of themes based on the Socio Ecological Model (SEM) domains.

**Table 5.**
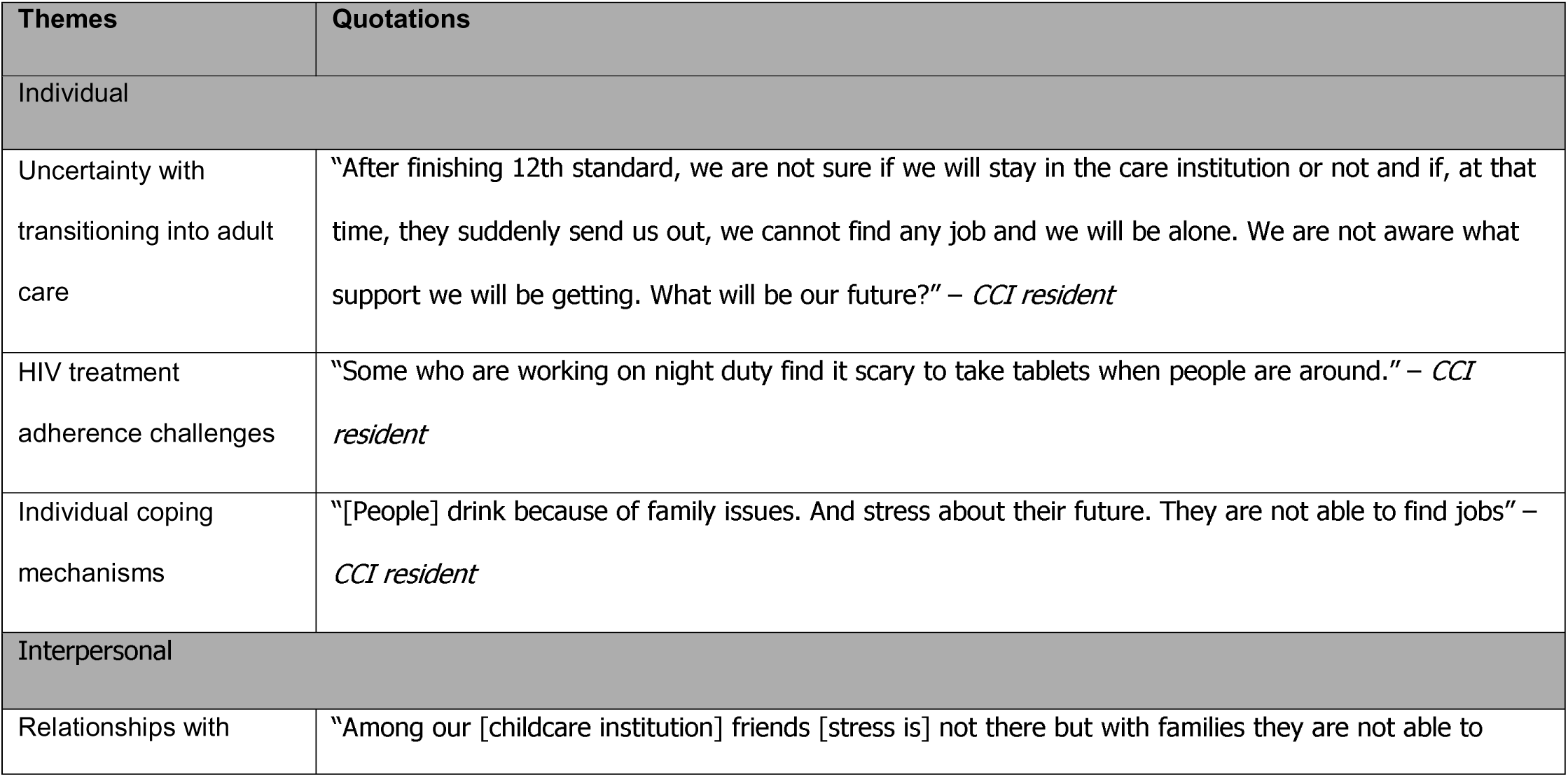
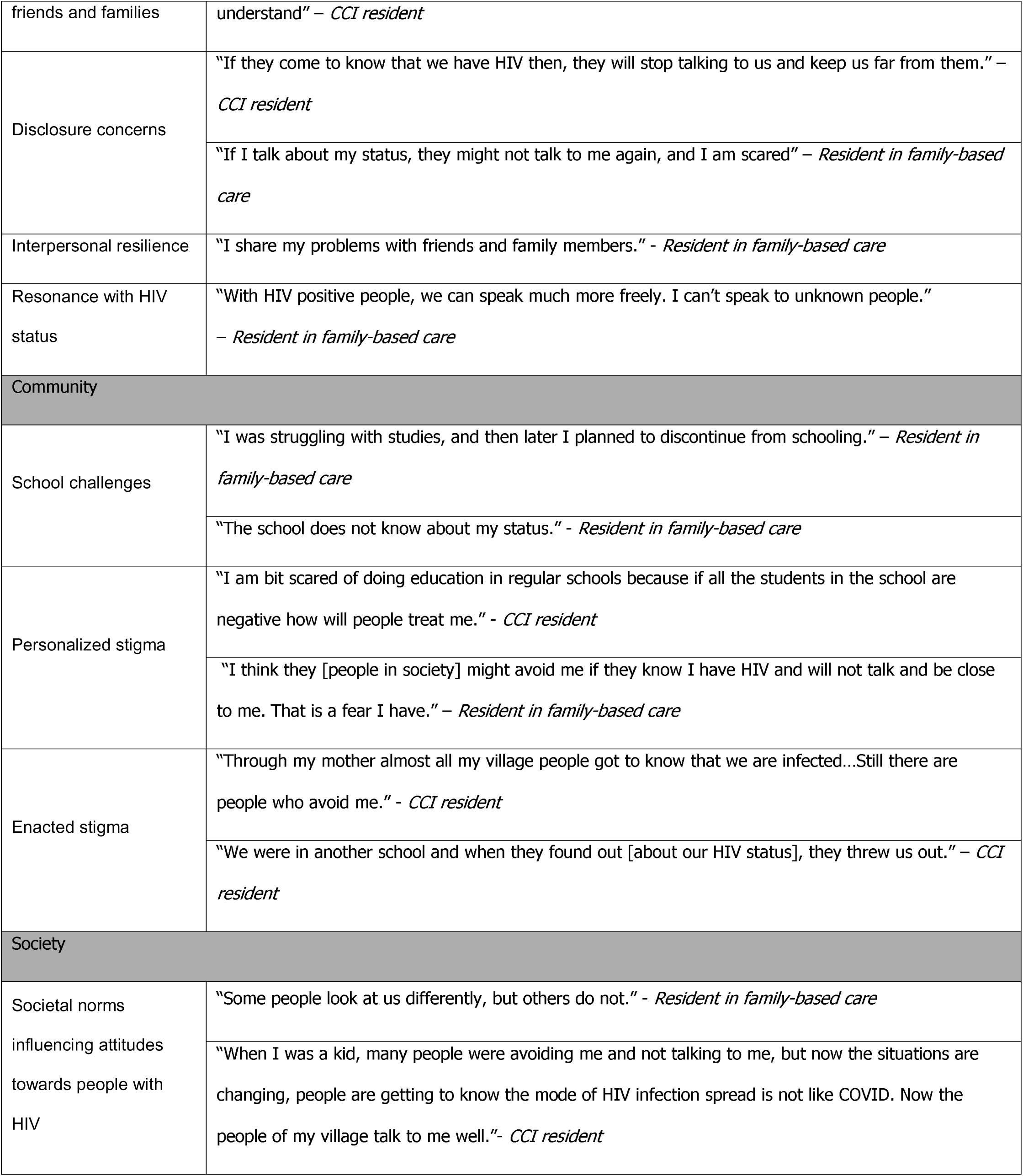
Peer participants’ quotations from focus group discussions.

### Individual factors

#### Uncertainty with transitioning into adult care

Participants’ concerns spanned across education, employment, ART adherence challenges, and future support after transitioning out of known care systems. (Table 5). APHIV who lived in childcare institutions expressed worries about navigating the health system after they left these institutions and in general described being underprepared for transition.

#### HIV treatment adherence challenges

While participants generally had high ART adherence, they identified situations where taking daily medications was challenging (for example, when at work, or when traveling), and found taking medications to be a stressful experience due to persistent concerns around inadvertent disclosure.

#### Individual coping mechanisms

Participants relied in part on individual coping mechanisms, such as distraction through hobbies and self-care activities. However, particularly among young adults who had transitioned from CCIs to independent living, participants described maladaptive coping mechanisms with substance use in order to deal with structural and psychosocial vulnerabilities including financial and housing insecurity, and poor social support.

### Interpersonal factors

#### Relationships with friends and families

Family support played a significant role as a protective interpersonal factor, whereas lack of family support was a contributor to symptoms of anxiety and/or depression. There were notable differences in family support between APHIV living in CCIs and those living in the community; the former were often double orphans and primarily relied on a sibling or other peers in the CCIs for support. They perceived their extended family members as lacking understanding, whereas APHIV living in the community received support from a parent (usually of shared HIV status) that they could easily confide in.

#### Disclosure concerns

Personalized stigma (a community factor) intersected with disclosure concerns (an interpersonal factor). Several participants expressed fears around others avoiding them or not wanting to talk to them or interact with them because of their HIV status. As a result, participants generally avoided disclosing their status to even people who knew them well.

#### Interpersonal resilience

Participants identified family and friends as key sources of emotional support, providing a comfortable space for sharing their feelings. Among family members, siblings, both with and without HIV, were often mentioned as individuals with whom participants spoke to and supported them the most. Some participants noted that family members that do not have HIV often struggle to understand their stress. Conversely, some participants expressed distrust towards their friends, fearing loss of confidentiality.

#### Resonance with HIV status

When discussing fellows who shared their HIV status, participants expressed that they felt no hesitation in sharing their challenges and mutual understanding. They often mentioned that it was easier to talk to someone who understood their situation compared to someone who did not. In particular, when it came to issues related to HIV health, such as hospital visits and obtaining ART, fellows proved to be vital sources of support.

### Community factors

#### School challenges

School was identified as a significant source of distress, stemming from various factors, including academic pressure, fear of others discovering their status, and lack of support to continue their education. Schools were also primary spaces where participants interacted with others without HIV, where they often faced challenges when their status was known, including having to move schools. Sources of anxiety included daily studies and exams, and financial support needed to continue their studies.

#### Personalized and enacted stigma

Participants expressed a strong belief that they would face rejection, isolation, or negative treatment from others due to their HIV status, leading to increased anxiety. This concern was particularly evident when discussing social interactions and general societal attitudes where participants feared that disclosure of their status could lead to social distancing or exclusion from events. These anxieties impacted their participation in activities and school attendance.

### Societal factors

#### Societal norms influencing attitudes towards people with HIV

Participants had more nuanced perceptions about societal and public attitudes towards people with HIV. While they observed a broader climate of stigma and negative views, they recognized that not all individuals embraced these attitudes. For example, one participant noted how, despite some community members avoiding them due to HIV status, others began to engage more positively as awareness about HIV transmission increased leading to greater acceptance. These experiences reflect that societal attitudes do not permeate monolithically into personal interactions, leading to more individualized perceptions of acceptance or rejection.

## Discussion

Within a contemporary context of widespread ART availability and accessibility in India, our study highlights unmet mental health vulnerabilities among APHIV despite achievement of treatment success with high rates of viral suppression. We identified these vulnerabilities within a peer support program, suggesting the need for tailored evidence-based interventions for APHIV that extend beyond viral suppression alone and address broader factors related to mental health and optimal transition into adult care and independent living.

Our estimates of prevalence of positive screens for CMD among APHIV have implications for youth in India regardless of achievement of HIV treatment success. Despite more than 90% of APHIV in our study being virally suppressed, more than half had positive screens for mild or greater severity of CMD. The prevalence of positive moderate and severe depression (14.6%) and moderate and severe anxiety (9.8%) in our cohort is comparable to studies among similar populations in other settings with high levels of viral suppression.^73–75^ For example, in a recent study in Thailand among a cohort of 100 young people with HIV, of whom 81.0% had undetectable viral loads, approximately 20% experienced positive screens for significant depression or anxiety.^75^ In particular, 7.0% had isolated depression, defined by a modified PHQ-9 for adolescents score of ≥10 (moderate severity or higher); 2% had isolated anxiety, defined by a Child Anxiety Related Disorder (SCARED) score of ≥25, thus indicating the presence of an anxiety disorder; 8% experienced comorbid anxiety and depression.^75^

Our study’s screening estimates are significant in that they signal the presence of a high burden of both mild and more severe CMD among APHIV who are highly engaged in HIV care. We found that double orphan status was significantly associated with increased odds of positive screens for anxiety, while other structural and psychosocial vulnerabilities, including school discontinuation and low resilience demonstrated directionality towards increased odds of positive screens for anxiety, depression, or at least one CMD. These vulnerabilities and other structural and psychosocial factors such as school exclusion, bereavement, and poverty have been significantly associated with CMD in other global studies among young people with HIV.^76–81^ Notably, studies across several settings suggest that structural and psychosocial vulnerabilities concentrate among young people with HIV who have not achieved viral suppression,^82–84^ lending support for a potentially higher burden of significant CMD in this group. One may anticipate that APHIV experiencing challenges with viral suppression and engagement in HIV care will have an even higher burden of moderate and above CMD severity.

Our study offers insights into distinct psychosocial factors related to mental health among sub-groups of APHIV, particularly those who live in CCIs and in family-based care. A substantial proportion of our participants lived in CCIs, with the rest living with a parent and/or extended families. In multivariable models, although there was no significant difference in association of positive screens for CMD between CCI-based APHIV versus others possibly owing to small sample sizes, qualitative exploration yielded narratives that highlight divergence around mental health stressors in these sub-groups. Family support, a well-known protective factor for CMD, was often lacking in APHIV living in CCIs compared to those living with a parent and/or extended family.^85–87^ Consistent with other studies,^88^ APHIV in CCIs received minimal family support and described strained relationships with extended family members as a source of stress, whereas for APHIV living with families, a parent with a shared HIV status, as well as other family members were sources of support, in addition to peers. Notably, the prospect of graduation from institutional care at age 18 years (a legal stipulation in India) and needing to navigate multiple transitions simultaneously, including transition to emerging adulthood and independent living with minimal preparation and supports, was a distinct stressor for adolescents under the age of 18 years living in CCIs.

Stigma narratives were consistent with other studies,^89,90^ with APHIV being highly concerned about societal and public attitudes towards people with HIV. Narratives around school discontinuation, a risk factor for CMD among youth, also centered around anticipatory and enacted stigma with examples of actual experiences of forced school exclusion due to inadvertent disclosure of HIV status. Strikingly, in our study, public attitudes, disclosure concerns, and anticipatory and enacted stigma did not necessarily translate to negative self-image among APHIV, indicating that external stigma was much more significant than internal stigma in this population. This may in part be related to receiving peer support that incorporates regular group discussions on shared experiences of stigma and related coping mechanisms.

The findings of our study underscore the need to develop and implement evidence-based and stigma-informed interventions for preventing and addressing CMD among APHIV that are also differentiated to address the unique vulnerabilities of sub-groups, such as orphans and those growing up in CCIs. Onset of significant CMD in adolescence, in particular depression and anxiety, confers a higher risk of recurrence in adulthood.^91–94^ For APHIV, addressing CMD early needs to be a priority as this has the potential to impact the trajectories of two independent, yet intersecting causes of ill-health and death–HIV and mental health disorders– across the life course in this population.

A significant health system gap to address CMD exists in India and other LMICs due to suboptimal availability of mental health professionals.^95^ Task-shifted stepped-care approaches for lay-person delivery of evidence-based interventions for CMD among adolescents have been successful in school-settings in India.^96–98^ However, as found in our study and others,^20,99,100^ youth with HIV encounter unique challenges in school settings and work settings, including school exclusion and significant concerns around disclosure. Therefore, spaces which these youth perceive as being “safe” may be better suited for task-shifted delivery of evidence-based interventions for CMD. Task-shifted approaches include leveraging existing ART counselors in public sector ART centers to screen for CMD and deliver evidence-based interventions. While high client volume at public sector ART centers may pose challenges to integrating task-shifted roles for mental health care delivery, youth-friendly models of HIV care led by trained youth and community members show promise globally, ^101–104^ and in Indian settings^105^ in addressing CMD among APHIV.

One consistent finding across studies, including ours, is the natural resonance around HIV status and lived experiences among APHIV, and consequently the preference for communication about challenges to peers. In various countries, peer supporters are part of national HIV programs, such as community treatment supporters for youth with HIV in Zimbabwe.^106,107^ Recent studies indicate that peer-delivered evidence-based mental health interventions, such as problem-solving therapy, can be integrated into peer support models to address CMD.^38,108,109^ Our study serves as a proof-of-concept that with partnership and investment, APHIV can not only provide peer support but also be trained to conduct screening for CMD among their peers. As the next step, peer-delivered interventions for CMD should be a focus of further development and research in India. Finally, developing and incorporating differentiated interventions that include additional components such as addressing externalized stigma, evidence-based ART adherence interventions,^110–112^ and evidence-based transition readiness tools for APHIV living in CCIs needs to be a key consideration for delivery of CMD interventions to AYPHIV.^113,114^

Our study has a few limitations. First, all participants in the cross-sectional survey were recipients of the I’mPossible peer mentorship intervention, which may have mitigated CMD risk while positively influencing self-image and viral suppression. However it is less likely that peer support alone would have improved significant CMD, since no CMD-specific interventions are presently incorporated in the I’mPossible Fellowship. Second, APHIV in this study were generally highly engaged in HIV care, and we were limited in assessing CMD among those who were under-engaged in HIV care or lost to follow up, suggesting that our findings may be an underestimation of the actual prevalence among all APHIV. Moreover, since our population included only those with perinatally acquired HIV, CMD patterns may be different in youth with behaviorally acquired HIV. Additional studies utilizing population-level sampling strategies are needed to estimate true prevalence of CMD among youth with HIV across regions in India. Third, our qualitative exploration did not thoroughly explore the association of gender with CMD. In general, women in India tend to have a higher prevalence of CMD influenced by gender-specific risk factors such as gender-based violence, socioeconomic inequality, and caregiving burdens, that are likely to be compounded for young women with HIV in India^101,102^ Fourth, the small sample size in our survey likely limited determination of statistical significance of correlates of CMD.

Our study also has several strengths. Few studies in India have utilized validated screening instruments such as the PHQ-9 or GAD-7^115^ to screen for CMD among APHIV; most studies thus far involve qualitative exploration or assess broader psychosocial outcomes such as quality of life.^32,33,116–119^ Our approach of a cross-sectional survey followed by a qualitative research approach provides a more rounded understanding of CMD and their determinants among APHIV. Additionally, participatory approaches involving trained APHIV as youth investigators engaged in this research is a distinct strength, and our study is among the first in India utilizing this approach.^44^ Responses to peer-delivered screening instruments are also less likely to be prone to social desirability bias, yielding more reliable estimates of positive screens for CMD.

## Conclusions

We conclude that the high burden of positive screens for CMD among APHIV despite achievement of HIV treatment success necessitates immediate and long-term investment to integrate mental health screening and intervention implementation across the pediatric-to-adult HIV care continuum for all adolescents with HIV. Implementation research that incorporates community-based participatory research principles is needed to inform both screening and delivery of evidence-based interventions for CMD among APHIV. Such investments need to be a high priority as they have the potential to not only shape HIV care outcomes but also yield lasting benefits beyond HIV across the life-course for young people with HIV.

## Data Availability

De-identified data used in this study may be accessed upon reasonable request to the
corresponding author.

## Acknowledgements

We thank our partner non-governmental organizations without whom our research would not have been possible.

## References

1. Adolescent HIV prevention [Internet]. UNICEF DATA. [cited 2024 Feb 29]. Available from: https://data.unicef.org/topic/hivaids/adolescents-young-people/

2. Young people and HIV. Available from: https://www.unaids.org/sites/default/files/media_asset/young-people-and-hiv_en.pdf

3. Arora A. Children and AIDS: Statistical update [Internet]. UNICEF DATA. 2017 [cited 2024 Jul 29]. Available from: https://data.unicef.org/resources/children-aids-statistical-update/

4. Slogrove AL, Sohn AH. The global epidemiology of adolescents living with HIV: time for more granular data to improve adolescent health outcomes. Curr Opin HIV AIDS. 2018 May;13(3):170–178. PMCID: PMC5929160

5. Christie D, Viner R. Adolescent development. BMJ. British Medical Journal Publishing Group; 2005 Feb 3;330(7486):301–304. PMID: 15695279

6. Jaworska N, MacQueen G. Adolescence as a unique developmental period. J Psychiatry Neurosci. 2015 Sep;40(5):291–293.

7. Gutgesell ME, Payne N. Issues of adolescent psychological development in the 21st century. Pediatr Rev. 2004 Mar;25(3):79–85. PMID: 14993515

8. Solmi M, Radua J, Olivola M, Croce E, Soardo L, Salazar de Pablo G, Il Shin J, Kirkbride JB, Jones P, Kim JH, Kim JY, Carvalho AF, Seeman MV, Correll CU, Fusar-Poli P. Age at onset of mental disorders worldwide: large-scale meta-analysis of 192 epidemiological studies. Mol Psychiatry. Nature Publishing Group; 2022 Jan;27(1):281–295.

9. Kessler RC, Berglund P, Demler O, Jin R, Merikangas KR, Walters EE. Lifetime prevalence and age-of-onset distributions of DSM-IV disorders in the National Comorbidity Survey Replication. Arch Gen Psychiatry. 2005 Jun;62(6):593–602. PMID: 15939837

10. Patton GC, Coffey C, Sawyer SM, Viner RM, Haller DM, Bose K, Vos T, Ferguson J, Mathers CD. Global patterns of mortality in young people: a systematic analysis of population health data. Lancet Lond Engl. 2009 Sep 12;374(9693):881–892. PMID: 19748397

11. Grossberg A, Rice T. Depression and Suicidal Behavior in Adolescents. Med Clin. Elsevier; 2023 Jan 1;107(1):169–182. PMID: 36402497

12. Lovero KL, Dos Santos PF, Come AX, Wainberg ML, Oquendo MA. Suicide in Global Mental Health. Curr Psychiatry Rep. 2023 Jun 1;25(6):255–262.

13. Vreeman RC, Scanlon ML, Marete I, Mwangi A, Inui TS, McAteer CI, Nyandiko WM. Characteristics of HIV-infected adolescents enrolled in a disclosure intervention trial in western Kenya. AIDS Care. Taylor & Francis; 2015 Nov 2;27(sup1):6–17. PMID: 26616121

14. Mellins CA, Malee KM. Understanding the mental health of youth living with perinatal HIV infection: lessons learned and current challenges. J Int AIDS Soc. 2013 Jun 18;16(1):18593. PMCID: PMC3687078

15. Ayano G, Demelash S, Abraha M, Tsegay L. The prevalence of depression among adolescent with HIV/AIDS: a systematic review and meta-analysis. AIDS Res Ther. 2021 Apr 27;18(1):23. PMCID: PMC8077927

16. Di Gennaro F, Marotta C, Ramirez L, Cardoso H, Alamo C, Cinturao V, Bavaro DF, Mahotas DC, Lazzari M, Fernando C, Chimundi N, Atzori A, Chaguruca I, Tognon F, Guambe Dos Anjos H, De Meneghi G, Tribie M, Del Greco F, Namarime E, Occa E, Putoto G, Pozniak A, Saracino A. High Prevalence of Mental Health Disorders in Adolescents and Youth Living with HIV: An Observational Study from Eight Health Services in Sofala Province, Mozambique. AIDS Patient Care STDs. Mary Ann Liebert, Inc., publishers; 2022 Apr;36(4):123–129.

17. Too EK, Abubakar A, Nasambu C, Koot HM, Cuijpers P, Newton CR, Nyongesa MK. Prevalence and factors associated with common mental disorders in young people living with HIV in sub-Saharan Africa: a systematic review. J Int AIDS Soc. 2021;24(S2):e25705.

18. Laurenzi CA, Skeen S, Gordon S, Akin-Olugbade O, Abrahams N, Bradshaw M, Brand A, du Toit S, Melendez-Torres GJ, Tomlinson M, Servili C, Dua T, Ross DA. Preventing mental health conditions in adolescents living with HIV: an urgent need for evidence. J Int AIDS Soc. 2020 Sep;23 Suppl 5(Suppl 5):e25556. PMCID: PMC7459172

19. Orth Z, Van Wyk B. Rethinking mental health wellness among adolescents living with HIV in the African context: An integrative review of mental wellness components. Front Psychol [Internet]. 2022 [cited 2024 Feb 29];13. Available from: https://www.frontiersin.org/journals/psychology/articles/10.3389/fpsyg.2022.955869

20. Kip EC, Udedi M, Kulisewa K, Go VF, Gaynes BN. Stigma and mental health challenges among adolescents living with HIV in selected adolescent-specific antiretroviral therapy clinics in Zomba District, Malawi. BMC Pediatr. 2022 May 6;22(1):253. PMCID: PMC9077887

21. Deike LG, Barreiro P, Reneses B. The new profile of psychiatric disorders in patients with HIV infection. AIDS Rev. 2023;25(1):41–53. PMID: 36952661

22. Boyes ME, Pantelic M, Casale M, Toska E, Newnham E, Cluver LD. Prospective associations between bullying victimisation, internalised stigma, and mental health in South African adolescents living with HIV. J Affect Disord. 2020 Nov 1;276:418–423. PMID: 32871672

23. Yehia BR, Stephens-Shield AJ, Momplaisir F, Taylor L, Gross R, Dubé B, Glanz K, Brady KA. Health Outcomes of HIV-Infected People with Mental Illness. AIDS Behav. 2015 Aug;19(8):1491–1500. PMCID: PMC4527875

24. Pence BW. The impact of mental health and traumatic life experiences on antiretroviral treatment outcomes for people living with HIV/AIDS. J Antimicrob Chemother. 2009 Apr 1;63(4):636–640.

25. Lang R, Hogan B, Zhu J, McArthur K, Lee J, Zandi P, Nestadt P, Silverberg MJ, Parcesepe AM, Cook JA, Gill MJ, Grelotti D, Closson K, Lima VD, Goulet J, Horberg MA, Gebo KA, Camoens RM, Rebeiro PF, Nijhawan AE, McGinnis K, Eron J, Althoff KN. The prevalence of mental health disorders in people with HIV and the effects on the HIV care continuum. AIDS. 2023 Feb 1;37(2):259.

26. Mayston R, Kinyanda E, Chishinga N, Prince M, Patel V. Mental disorder and the outcome of HIV/AIDS in low-income and middle-income countries: a systematic review. AIDS. 2012 Dec;26:S117.

27. Jahn A, Floyd S, Crampin AC, Mwaungulu F, Mvula H, Munthali F, McGrath N, Mwafilaso J, Mwinuka V, Mangongo B, Fine PE, Zaba B, Glynn JR. Population-level effect of HIV on adult mortality and early evidence of reversal after introduction of antiretroviral therapy in Malawi. Lancet. 2008 May 10;371(9624):1603–1611. PMCID: PMC2387197

28. Oguntibeju OO. Quality of life of people living with HIV and AIDS and antiretroviral therapy. HIVAIDS Auckl NZ. Dove Press; 2012;4:117. PMID: 22893751

29. Zhao Y, Wu Z, McGoogan JM, Shi CX, Li A, Dou Z, Ma Y, Qin Q, Brookmeyer R, Detels R, Montaner JSG. Immediate Antiretroviral Therapy Decreases Mortality Among Patients With High CD4 Counts in China: A Nationwide, Retrospective Cohort Study. Clin Infect Dis Off Publ Infect Dis Soc Am. 2018 Feb 10;66(5):727–734. PMCID: PMC5850406

30. Shao Y, Williamson C. The HIV-1 Epidemic: Low- to Middle-Income Countries. Cold Spring Harb Perspect Med. 2012 Mar;2(3):a007187. PMCID: PMC3282497

31. Wainberg ML, Scorza P, Shultz JM, Helpman L, Mootz JJ, Johnson KA, Neria Y, Bradford JME, Oquendo MA, Arbuckle MR. Challenges and Opportunities in Global Mental Health: a Research-to-Practice Perspective. Curr Psychiatry Rep. 2017 May;19(5):28. PMCID: PMC5553319

32. Verma A, Kota KK, Bangar S, Rahane G, Yenbhar N, Sahay S. Emotional distress among adolescents living with perinatal HIV in India: examining predictors and their mediating and moderating effects. Child Adolesc Psychiatry Ment Health. 2023 Mar 15;17(1):40. PMCID: PMC10018828

33. Gupta R, Shringi S, Mahajan V, G V, Srivastava K. A Study of Psychological Impact of Diagnosis of HIV in Children and Adolescents in Indian Population. HIVAIDS Res Treat - Open J. 2015 Feb 15;1(1):16–20.

34. Youth | National AIDS Control Organization | MoHFW | GoI [Internet]. [cited 2024 Feb 29]. Available from: https://naco.gov.in/youth

35. The path that ends AIDS: UNAIDS Global AIDS Update 2023 [Internet]. [cited 2024 Feb 29]. Available from: https://www.unaids.org/en/resources/documents/2023/global-aids-update-2023

36. HIV/AIDS Data Hub for the Asia Pacific [Internet]. HIV/AIDS Data Hub for the Asia Pacific. [cited 2024 Sep 27]. Available from: https://www.aidsdatahub.org

37. Bhana A, Kreniske P, Pather A, Abas MA, Mellins CA. Interventions to address the mental health of adolescents and young adults living with or affected by HIV: state of the evidence. J Int AIDS Soc. 2021 Jun 24;24(Suppl 2):e25713. PMCID: PMC8222850

38. Simms V, Weiss HA, Chinoda S, Mutsinze A, Bernays S, Verhey R, Wogrin C, Apollo T, Mugurungi O, Sithole D, Chibanda D, Willis N. Peer-led counselling with problem discussion therapy for adolescents living with HIV in Zimbabwe: A cluster-randomised trial. PLOS Med. Public Library of Science; 2022 Jan 5;19(1):e1003887.

39. Bhana A, McKay MM, Mellins C, Petersen I, Bell C. Family-based HIV prevention and intervention services for youth living in poverty-affected contexts: the CHAMP model of collaborative, evidence-informed programme development. J Int AIDS Soc. 2010 Jun 23;13(Suppl 2):S8. PMCID: PMC2890977

40. Betancourt TS, Ng LC, Kirk CM, Brennan RT, Beardslee WR, Stulac S, Mushashi C, Nduwimana E, Mukunzi S, Nyirandagijimana B, Kalisa G, Cyamatare FR, Sezibera V. Family-Based Promotion of Mental Health in Children Affected by HIV: A Pilot Randomized Controlled Trial. J Child Psychol Psychiatry. 2017 Aug;58(8):922–930. PMCID: PMC5730278

41. Webb L, Perry-Parrish C, Ellen J, Sibinga E. Mindfulness instruction for HIV-infected youth: a randomized controlled trial. AIDS Care. 2018 Jun;30(6):688–695. PMCID: PMC5987527

42. Cavazos-Rehg P, Byansi W, Xu C, Nabunya P, Bahar OS, Borodovsky J, Kasson E, Anako N, Mellins C, Damulira C, Neilands T, Ssewamala FM. The Impact of a Family-Based Economic Intervention on the Mental Health of HIV-Infected Adolescents in Uganda: Results From Suubi + Adherence. J Adolesc Health Off Publ Soc Adolesc Med. 2021 Apr;68(4):742–749. PMCID: PMC7987910

43. Raj M, Sannigrahi S, Sidduramu R, Seenappa B, Reddy L, Sharma A, Kumar S, Solomon S, Ganapathi L, Shet A. Building resilience through peer mentorship among adolescents and young adults with perinatal HIV: The impact of the I’mpossible Fellowship intervention in India. Abstr -AIDS-2024-06152. 25th International AIDS Conference, Munich, Germany; 2024.

44. Sannigrahi S, Seenappa B, Lakshmikanth P, Reddy S, Filian K, Raj MB, Ganapathi L, Shet A. Partnering for Progress: Lessons Learned From Mental Health Assessment for Youth Living With HIV in India Through Community-Based Participatory Research. J Particip Res Methods [Internet]. 2024 Oct 9 [cited 2024 Dec 25];5(3). Available from: https://jprm.scholasticahq.com/article/117611-partnering-for-progress-lessons-learned-from-mental-health-assessment-for-youth-living-with-hiv-in-india-through-community-based-participatory-resear/stats/all/pageviews

45. Saal W, Thomas A, Mangqalaza H, Laurenzi C, Kelly J, Toska E. Adapting the CYRM-R for use among adolescent girls and young mothers affected by HIV in South Africa Cognitive interview methodology and results Adapting the CYRM-R for use among adolescent girls and young mothers affected by HIV in South Africa. Cognitive interview methodology and results. 2022.

46. Kaunda-Khangamwa BN, Maposa I, Dambe R, Malisita K, Mtagalume E, Chigaru L, Munthali A, Chipeta E, Phiri S, Manderson L. Validating a Child Youth Resilience Measurement (CYRM-28) for Adolescents Living With HIV (ALHIV) in Urban Malawi. Front Psychol [Internet]. 2020 [cited 2024 Feb 28];11. Available from: https://www.frontiersin.org/journals/psychology/articles/10.3389/fpsyg.2020.01896

47. Singh K, Bandyopadhyay S, Raina M. Validation of the Child and Youth Resilience Measure-28 (CYRM-28) in India. J Indian Assoc Child Adolesc Ment Health. SAGE Publications; 2022 Jul 1;18(3):218–225.

48. De Man J, Absetz P, Sathish T, Desloge A, Haregu T, Oldenburg B, Johnson LCM, Thankappan KR, Williams ED. Are the PHQ-9 and GAD-7 Suitable for Use in India? A Psychometric Analysis. Front Psychol. 2021 May 13;12:676398. PMCID: PMC8155718

49. Ungar M. Systemic resilience: principles and processes for a science of change in contexts of adversity. Ecol Soc. 2018;23(4):art34.

50. McDonald-Harker C, Drolet JL, Sehgal A, Brown MRG, Silverstone PH, Brett-MacLean P, Agyapong VIO. Social-Ecological Factors Associated With Higher Levels of Resilience in Children and Youth After Disaster: The Importance of Caregiver and Peer Support. Front Public Health. 2021 Jul 29;9:682634. PMCID: PMC8358203

51. Kroenke K, Spitzer RL, Williams JBW. The PHQ-9. J Gen Intern Med. 2001 Sep;16(9):606–613. PMCID: PMC1495268

52. Johnson SU, Ulvenes PG, Øktedalen T, Hoffart A. Psychometric Properties of the General Anxiety Disorder 7-Item (GAD-7) Scale in a Heterogeneous Psychiatric Sample. Front Psychol [Internet]. Frontiers; 2019 Aug 6 [cited 2024 Sep 27];10. Available from: https://www.frontiersin.org/journals/psychology/articles/10.3389/fpsyg.2019.01713/full

53. Marbaniang I, Borse R, Sangle S, Kinikar A, Chavan A, Nimkar S, Suryavanshi N, Mave V. Development of shortened HIV-related stigma scales for young people living with HIV and young people affected by HIV in India. Health Qual Life Outcomes. 2022 Jul 31;20(1):119.

54. Reinius M, Wettergren L, Wiklander M, Svedhem V, Ekström AM, Eriksson LE. Development of a 12-item short version of the HIV stigma scale. Health Qual Life Outcomes. 2017 May 30;15(1):115.

55. Saine ME, Moore TM, Szymczak JE, Bamford LP, Barg FK, Mitra N, Schnittker J, Holmes JH, Lo Re V. Validation of a modified Berger HIV stigma scale for use among patients with hepatitis C virus (HCV) infection. PLoS ONE. 2020 Feb 5;15(2):e0228471. PMCID: PMC7001940

56. Berger BE, Ferrans CE, Lashley FR. Measuring stigma in people with HIV: psychometric assessment of the HIV stigma scale. Res Nurs Health. 2001 Dec;24(6):518–529. PMID: 11746080

57. Earnshaw VA, Smith LR, Chaudoir SR, Amico KR, Copenhaver MM. HIV Stigma Mechanisms and Well-Being among PLWH: A Test of the HIV Stigma Framework. AIDS Behav. 2013 Jun;17(5):1785–1795. PMCID: PMC3664141

58. Baral S, Logie CH, Grosso A, Wirtz AL, Beyrer C. Modified social ecological model: a tool to guide the assessment of the risks and risk contexts of HIV epidemics. BMC Public Health. 2013 May 17;13(1):482.

59. Spitzer RL, Kroenke K, Williams JBW, Löwe B. Generalized Anxiety Disorder 7 [Internet]. 2011 [cited 2024 Feb 28]. Available from: https://doi.apa.org/doi/10.1037/t02591-000

60. Solomon SS, Mehta SH, McFall AM, Srikrishnan AK, Saravanan S, Laeyendecker O, Balakrishnan P, Celentano DD, Solomon S, Lucas GM. Community viral load, antiretroviral therapy coverage, and HIV incidence in India: A cross-sectional, comparative evaluation study. Lancet HIV. 2016 Apr;3(4):e183–e190. PMCID: PMC4863069

61. Thorman J, Björkman P, Sasinovich S, Tesfaye F, Mulleta D, Medstrand P, Reepalu A. Performance of Galectin-9 for Identification of HIV Viremia in Adults Receiving Antiretroviral Therapy in a Resource-Limited Setting. J Acquir Immune Defic Syndr 1999. 2023 Jul 1;93(3):244–250. PMID: 36961948

62. Seedat S, Scott KM, Angermeyer MC, Berglund P, Bromet EJ, Brugha TS, Demyttenaere K, de Girolamo G, Haro JM, Jin R, Karam EG, Kovess-Masfety V, Levinson D, Mora MEM, Ono Y, Ormel J, Pennell BE, Posada-Villa J, Sampson NA, Williams D, Kessler RC. Cross-national associations between gender and mental disorders in the WHO World Mental Health Surveys. Arch Gen Psychiatry. 2009 Jul;66(7):785–795. PMCID: PMC2810067

63. Shiferaw G, Bacha L, Tsegaye D. Prevalence of Depression and Its Associated Factors among Orphan Children in Orphanages in Ilu Abba Bor Zone, South West Ethiopia. Psychiatry J. 2018;2018:6865085. PMCID: PMC6205311

64. Weich S, Twigg L, Lewis G. Rural/non-rural differences in rates of common mental disorders in Britain: prospective multilevel cohort study. Br J Psychiatry J Ment Sci. 2006 Jan;188:51–57. PMID: 16388070

65. McCloud T, Kamenov S, Callender C, Lewis G, Lewis G. The association between higher education attendance and common mental health problems among young people in England: evidence from two population-based cohorts. Lancet Public Health. 2023 Oct;8(10):e811–e819. PMID: 37777290

66. Evans L, Lund C, Massazza A, Weir H, Fuhr DC. The impact of employment programs on common mental disorders: A systematic review. Int J Soc Psychiatry. 2022 Nov;68(7):1315–1323. PMCID: PMC9548920

67. Nguyen N, Lovero KL, Falcao J, Brittain K, Zerbe A, Wilson IB, Kapogiannis B, Pimentel De Gusmao E, Vitale M, Couto A, Simione TB, Abrams EJ, Mellins CA. Mental health and ART adherence among adolescents living with HIV in Mozambique. AIDS Care. 2023 Feb;35(2):182–190. PMCID: PMC10243515

68. Da W, Li X, Qiao S, Zhou Y, Shen Z. Antiretroviral therapy and mental health among people living with HIV/AIDS in China. Psychol Health Med. 2020 Jan;25(1):45–52. PMCID: PMC6842406

69. Haas AD, Lienhard R, Didden C, Cornell M, Folb N, Boshomane TMG, Salazar-Vizcaya L, Ruffieux Y, Nyakato P, Wettstein AE, Tlali M, Davies MA, von Groote P, Wainberg M, Egger M, Maartens G, Joska JA. Mental Health, ART Adherence, and Viral Suppression Among Adolescents and Adults Living with HIV in South Africa: A Cohort Study. AIDS Behav. 2023 Jun;27(6):1849–1861. PMCID: PMC10149479

70. Srivastava K. Positive mental health and its relationship with resilience. Ind Psychiatry J. 2011;20(2):75–76. PMCID: PMC3530291

71. Weitzel EC, Löbner M, Glaesmer H, Hinz A, Zeynalova S, Henger S, Engel C, Reyes N, Wirkner K, Löffler M, Riedel-Heller SG. The Association of Resilience with Mental Health in a Large Population-Based Sample (LIFE-Adult-Study). Int J Environ Res Public Health. 2022 Nov 29;19(23):15944. PMCID: PMC9740913

72. Färber F, Rosendahl J. The Association Between Resilience and Mental Health in the Somatically Ill. Dtsch Arzteblatt Int. 2018 Sep 21;115(38):621–627. PMCID: PMC6218704

73. Le Prevost M, Arenas-Pinto A, Melvin D, Parrott F, Foster C, Ford D, Evangeli M, Winston A, Sturgeon K, Rowson K, Gibb DM, Judd A, on behalf of the Adolescents and Adults Living with Perinatal HIV (AALPHI) Steering Committee. Anxiety and depression symptoms in young people with perinatally acquired HIV and HIV affected young people in England. AIDS Care. 2018 Aug 3;30(8):1040–1049.

74. Kemigisha E, Zanoni B, Bruce K, Menjivar R, Kadengye D, Atwine D, Rukundo GZ. Prevalence of depressive symptoms and associated factors among adolescents living with HIV/AIDS in South Western Uganda. AIDS Care. Taylor & Francis; 2019 Oct 3;31(10):1297–1303. PMID: 30621430

75. Chantaratin S, Trimetha K, Werarak P, Lapphra K, Maleesatharn A, Rungmaitree S, Wittawatmongkol O, Phongsamart W, Kongstan N, Khumcha B, Chokephaibulkit K. Depression and Anxiety in Youth and Young Adults Living with HIV: Frequency and Associated Factors in Thai Setting. J Int Assoc Provid AIDS Care. 2022 May 17;21:23259582221101811. PMCID: PMC9121500

76. Chuang SP, Wu JYW, Wang CS. Resilience and Quality of Life in People with Mental Illness: A Systematic Review and Meta-Analysis. Neuropsychiatr Dis Treat. 2023 Mar 4;19:507–514. PMCID: PMC9994666

77. Dulin AJ, Dale SK, Earnshaw VA, Fava JL, Mugavero MJ, Napravnik S, Hogan JW, Carey MP, Howe CJ. Resilience and HIV: A review of the definition and study of resilience. AIDS Care. 2018 Aug;30(SUP5):S6–S17. PMCID: PMC6436992

78. Wortsman B, Brice H, Capani A, Ball MC, Zinszer B, Tanoh F, Akpé H, Ogan A, Wolf S, Jasińska K. Risk and resilience factors for primary school dropout in Côte d’Ivoire. J Appl Dev Psychol. 2024 May 1;92:101654.

79. Cerniglia L, Cimino S, Ballarotto G, Monniello G. Parental Loss During Childhood and Outcomes on Adolescents’ Psychological Profiles: A Longitudinal Study. Curr Psychol. 2014 Dec 1;33(4):545–556.

80. Stikkelbroek Y, Bodden DHM, Reitz E, Vollebergh WAM, van Baar AL. Mental health of adolescents before and after the death of a parent or sibling. Eur Child Adolesc Psychiatry. 2016 Jan 1;25(1):49–59.

81. Tebeka S, Hoertel N, Dubertret C, Le Strat Y. Parental Divorce or Death During Childhood and Adolescence and Its Association With Mental Health. J Nerv Ment Dis. 2016 Sep;204(9):678.

82. Ndongo FA, Kana R, Nono MT, Noah JPYA, Ndzie P, Tejiokem MC, Biheng EH, Ndie J, Nkoa TA, Ketchaji A, Ngako JN, Penda CI, Bissek ACZK, Ndombo POK, Hawa HM, Msellati P, Lallemant M, Faye A. Association between mental disorders with detectable viral load and poor adherence to antiretroviral therapy among adolescents infected with Human Immunodeficiency Virus on follow-up at Chantal Biya Foundation, Cameroon. J Epidemiol Popul Health. 2024 Apr 1;72(2):202193.

83. Chenneville T, Gabbidon K, Lynn C, Rodriguez C. Psychological factors related to resilience and vulnerability among youth with HIV in an integrated care setting. AIDS Care. 2018;30(sup4):5–11. PMID: 30632781

84. Crowley T, van der Merwe AS, Esterhuizen T, Skinner D. Resilience of adolescents living with HIV in the Cape Metropole of the Western Cape. AIDS Care. 2022 Sep;34(9):1103–1110. PMID: 34378464

85. Chen P, Harris KM. Association of Positive Family Relationships With Mental Health Trajectories From Adolescence to Midlife. JAMA Pediatr. 2019 Dec;173(12):e193336. PMCID: PMC6784807

86. Wowolo G, Cao W, Bosomtwe D, Enimil A, Tarantino N, Barker DH, Galárraga O. The Impact of Different Parental Figures of Adolescents Living With HIV: An Evaluation of Family Structures, Perceived HIV Related Stigma, and Opportunities for Social Support. Front Public Health. 2022 Mar 24;10:647960. PMCID: PMC8987121

87. Betancourt TS, Meyers-Ohki SE, Charrow A, Hansen N. Mental Health and Resilience in HIV/AIDS-Affected Children: A Review of the Literature and Recommendations for Future Research. J Child Psychol Psychiatry. 2013 Apr;54(4):423–444. PMCID: PMC3656822

88. Nestadt DF, Alicea S, Petersen I, John S, Myeza NP, Nicholas SW, Cohen LG, Holst H, Bhana A, McKay MM, Abrams EJ, Mellins CA. HIV+ and HIV− youth living in group homes in South Africa need more psychosocial support. Vulnerable Child Youth Stud. 2013 Jul 1;8(3):195–205. PMCID: PMC3769796

89. Armoon B, Fleury MJ, Bayat AH, Fakhri Y, Higgs P, Moghaddam LF, Gonabadi-Nezhad L. HIV related stigma associated with social support, alcohol use disorders, depression, anxiety, and suicidal ideation among people living with HIV: a systematic review and meta-analysis. Int J Ment Health Syst. 2022 Mar 4;16(1):17.

90. MacLean JR, Wetherall K. The Association between HIV-Stigma and Depressive Symptoms among People Living with HIV/AIDS: A Systematic Review of Studies Conducted in South Africa. J Affect Disord. 2021 May 15;287:125–137.

91. Naicker K, Galambos NL, Zeng Y, Senthilselvan A, Colman I. Social, demographic, and health outcomes in the 10 years following adolescent depression. J Adolesc Health Off Publ Soc Adolesc Med. 2013 May;52(5):533–538. PMID: 23499382

92. Pine DS, Cohen P, Gurley D, Brook J, Ma Y. The Risk for Early-Adulthood Anxiety and Depressive Disorders in Adolescents With Anxiety and Depressive Disorders. Arch Gen Psychiatry. 1998 Jan 1;55(1):56–64.

93. Fergusson DM, Boden JM, Horwood LJ. Recurrence of major depression in adolescence and early adulthood, and later mental health, educational and economic outcomes. Br J Psychiatry. 2007 Oct;191(4):335–342.

94. Bruce SE, Yonkers KA, Otto MW, Eisen JL, Weisberg RB, Pagano M, Shea MT, Keller MB. Influence of psychiatric comorbidity on recovery and recurrence in generalized anxiety disorder, social phobia, and panic disorder: a 12-year prospective study. Am J Psychiatry. 2005 Jun;162(6):1179–1187. PMCID: PMC3272761

95. Weaver LJ, Karasz A, Muralidhar K, Jaykrishna P, Krupp K, Madhivanan P. Will increasing access to mental health treatment close India’s mental health gap? SSM - Ment Health. 2023 Dec 1;3:100184.

96. Michelson D, Malik K, Parikh R, Weiss HA, Doyle AM, Bhat B, Sahu R, Chilhate B, Mathur S, Krishna M, Sharma R, Sudhir P, King M, Cuijpers P, Chorpita B, Fairburn CG, Patel V. Effectiveness of a brief lay counsellor-delivered, problem-solving intervention for adolescent mental health problems in urban, low-income schools in India: a randomised controlled trial. Lancet Child Adolesc Health. 2020 Aug;4(8):571–582. PMCID: PMC7386943

97. Michelson D, Malik K, Krishna M, Sharma R, Mathur S, Bhat B, Parikh R, Roy K, Joshi A, Sahu R, Chilhate B, Boustani M, Cuijpers P, Chorpita B, Fairburn CG, Patel V. Development of a transdiagnostic, low-intensity, psychological intervention for common adolescent mental health problems in Indian secondary schools. Behav Res Ther. 2020 Jul;130:103439. PMCID: PMC7322400

98. Gellatly R, Knudsen K, Boustani MM, Michelson D, Malik K, Mathur S, Nair P, Patel V, Chorpita BF. A qualitative analysis of collaborative efforts to build a school-based intervention for multiple common adolescent mental health difficulties in India. Front Psychiatry [Internet]. Frontiers; 2022 Nov 24 [cited 2024 Sep 9];13. Available from: https://www.frontiersin.org/journals/psychiatry/articles/10.3389/fpsyt.2022.1038259/full

99. Robinson A, Cooney A, Fassbender C, McGovern DP. Examining the Relationship Between HIV-Related Stigma and the Health and Wellbeing of Children and Adolescents Living with HIV: A Systematic Review. AIDS Behav. 2023;27(9):3133–3149. PMCID: PMC10386953

100. Kimera E, Vindevogel S, Reynaert D, Justice KM, Rubaihayo J, De Maeyer J, Engelen AM, Musanje K, Bilsen J. Experiences and effects of HIV-related stigma among youth living with HIV/AIDS in Western Uganda: A photovoice study. PLoS ONE. 2020 Apr 24;15(4):e0232359. PMCID: PMC7182188

101. Reif LK, Rivera VR, Bertrand R, Belizaire ME, Joseph JMB, Louis B, Joseph B, Anglade B, Seo G, Severe P, Rouzier V, Pape JW, Fitzgerald DW, McNairy ML. “FANMI”: A Promising Differentiated Model of HIV Care for Adolescents in Haiti. JAIDS J Acquir Immune Defic Syndr. 2019 Sep 1;82(1):e11.

102. Mavhu W, Willis N, Mufuka J, Bernays S, Tshuma M, Mangenah C, Maheswaran H, Mangezi W, Apollo T, Araya R, Weiss HA, Cowan FM. Effect of a differentiated service delivery model on virological failure in adolescents with HIV in Zimbabwe (Zvandiri): a cluster-randomised controlled trial. Lancet Glob Health. 2020 Feb 1;8(2):e264–e275.

103. Ssewamala FM, Dvalishvili D, Mellins CA, Geng EH, Makumbi F, Neilands TB, McKay M, Damulira C, Nabunya P, Bahar OS, Nakigozi G, Kigozi G, Byansi W, Mukasa M, Namuwonge F. The long-term effects of a family based economic empowerment intervention (Suubi+Adherence) on suppression of HIV viral loads among adolescents living with HIV in southern Uganda: Findings from 5-year cluster randomized trial. PLOS ONE. Public Library of Science; 2020 Feb 10;15(2):e0228370.

104. Ahmed CV, Doyle R, Gallagher D, Imoohi O, Ofoegbu U, Wright R, Yore MA, Brooks MJ, Flores DD, Lowenthal ED, Rice BM, Buttenheim AM. A Systematic Review of Peer Support Interventions for Adolescents Living with HIV in Sub-Saharan Africa. AIDS Patient Care STDs. Mary Ann Liebert, Inc., publishers; 2023 Nov;37(11):535–559.

105. Arumugam V. Improving health outcomes among adolescents living with HIV: comprehensive service delivery contributes to nearly universal viral suppression among ALHIV in Imphal, India [Internet]. IAS Programme. [cited 2024 Sep 13]. Available from: https://programme.ias2023.org/mobile/abstract/abstractlist

106. Bernays S, Tshuma M, Willis N, Mvududu K, Chikeya A, Mufuka J, Cowan F, Mavhu W. Scaling up peer□led community□based differentiated support for adolescents living with HIV: keeping the needs of youth peer supporters in mind to sustain success. J Int AIDS Soc. 2020 Aug 31;23(Suppl 5):e25570. PMCID: PMC7459167

107. Tapera T, Willis N, Madzeke K, Napei T, Mawodzeke M, Chamoko S, Mutsinze A, Zvirawa T, Dupwa B, Mangombe A, Chimwaza A, Makoni TM, Mandewo W, Senkoro M, Owiti P, Tripathy JP, Kumar AMV. Effects of a Peer-Led Intervention on HIV Care Continuum Outcomes Among Contacts of Children, Adolescents, and Young Adults Living With HIV in Zimbabwe. Glob Health Sci Pract. 2019 Dec 23;7(4):575–584. PMCID: PMC6927836

108. Puschner B, Repper J, Mahlke C, Nixdorf R, Basangwa D, Nakku J, Ryan G, Baillie D, Shamba D, Ramesh M, Moran G, Lachmann M, Kalha J, Pathare S, Müller-Stierlin A, Slade M. Using Peer Support in Developing Empowering Mental Health Services (UPSIDES): Background, Rationale and Methodology. Ann Glob Health. 85(1):53. PMCID: PMC6634474

109. Giusto A, Vander Missen MR, Kosgei G, Njiriri F, Puffer E, Kamaru Kwobah E, Barasa J, Turissini M, Rasmussen J, Ott M, Binayo J, Rono W, Jaguga F. Peer-delivered Problem-solving Therapy for Adolescent Mental Health in Kenya: Adaptation for Context and Training of Peer-counselors. Res Child Adolesc Psychopathol. 2023 May 23;1–14. PMCID: PMC10203666

110. Safren SA, Otto MW, Worth JL. Life-steps: Applying cognitive behavioral therapy to HIV medication adherence. Cogn Behav Pract. 1999 Sep 1;6(4):332–341.

111. Shaw S, Amico KR. Antiretroviral Therapy Adherence Enhancing Interventions for Adolescents and Young Adults 13–24 Years of Age: A Review of the Evidence Base. JAIDS J Acquir Immune Defic Syndr. 2016 Aug 1;72(4):387.

112. Reif LK, Abrams EJ, Arpadi S, Elul B, McNairy ML, Fitzgerald DW, Kuhn L. Interventions to Improve Antiretroviral Therapy Adherence Among Adolescents and Youth in Low- and Middle-Income Countries: A Systematic Review 2015– 2019. AIDS Behav. 2020 Oct 1;24(10):2797–2810.

113. Dahourou DL, Gautier-Lafaye C, Teasdale CA, Renner L, Yotebieng M, Desmonde S, Ayaya S, Davies MA, Leroy V. Transition from paediatric to adult care of adolescents living with HIV in sub-Saharan Africa: challenges, youth-friendly models, and outcomes. J Int AIDS Soc. 2017 May 16;20(Suppl 3):21528. PMCID: PMC5577723

114. Ritchwood TD, Malo V, Jones C, Metzger IW, Atujuna M, Marcus R, Conserve DF, Handler L, Bekker LG. Healthcare retention and clinical outcomes among adolescents living with HIV after transition from pediatric to adult care: a systematic review. BMC Public Health. 2020 Aug 3;20(1):1195.

115. SG PK, G AK, SP R, V VS, Dandona R. A comparative assessment of generalized anxiety, conduct and peer relationship problems among AIDS and other orphaned children in India. BMC Psychiatry. 2016 Sep 21;16(1):330.

116. Perumbil Pathrose S, Washington RG, Washington M, Raj M, Sreenath K, Sudhesh NT, He S, Ramjan L. Quality of life of children and adolescents living with HIV in India: A systematic review and meta-analysis. Vulnerable Child Youth Stud. United Kingdom: Taylor & Francis; 2024;19(1):103–123.

117. Das S, Mukherjee A, Lodha R, Vatsa M. Quality of life and psychosocial functioning of HIV infected children. Indian J Pediatr. 2010 Jun 1;77(6):633–637.

118. Gopakumar KG, Bhat KG, Baliga S, Joseph N, Mohan N, Shetty AK. Impact of care at foster homes on the health-related quality of life of HIV-infected children and adolescents: a cross-sectional study from India. Qual Life Res. 2018 Apr 1;27(4):871–877.

119. Banerjee T, Pensi T, Banerjee D. HRQoL in HIV-infected children using PedsQL 4.0 and comparison with uninfected children. Qual Life Res Int J Qual Life Asp Treat Care Rehabil. 2010 Aug;19(6):803–812. PMID: 20383660

